# Task-dependent model selection for structured extraction from multilingual non-English clinical records

**DOI:** 10.64898/2026.09.03.26362149

**Authors:** Temirgali Aimyshev, Iliyar Arupzhanov, Gulnur Zhakhina, Aidana Rakhmankulova, Saltanat Smagul, Abduzhappar Gaipov

## Abstract

**Objective:** To evaluate task-dependent selection of local and cloud models for structured extraction from multilingual non-English stroke discharge summaries, distinguishing entity detection, record assembly, and raw-document processing.

**Methods:** This retrospective system evaluation compared multilingual encoders, locally fine-tuned Qwen3-4B models, and zero-shot GPT-5.5. Primary test cohorts comprised 332 section cases, 149 medication cases with 2,475 reference records after identity-based exclusions, and 191 laboratory cases. Outcomes were section-span F1, drug-name F1, normalized seven-field record recovery, test-name F1, and laboratory quintuple F1. Paired human comparisons used a common second-annotator reference on 50 cases per task. Saved cascade outputs were compared with curated-section controls using document-bootstrap intervals.

**Results:** Section F1 was 0.919 for the encoder, 0.926 for GPT-5.5, and 0.932 for Qwen. GPT-5.5 led medication detection (0.966 versus 0.940 for Qwen), whereas Qwen led normalized medication recovery (0.381 versus 0.311; difference 0.070, 95% CI 0.026–0.114) and laboratory quintuple F1 (0.892 versus 0.822). With matched inference stacks, medication recovery declined from 0.377 on curated inputs to 0.204 for single-window and 0.246 for all-block cascades; quintuple F1 on the original laboratory cohort declined from 0.898 to 0.811. Corpus-wide extraction processed 193,101 summaries in 13.3 H100 GPU-hours.

**Conclusion:** Local models achieved strong detection performance without frontier-scale inference in this setting. Fine-tuned local LLMs were advantageous for record assembly under the evaluated recipes; annotation conventions, normalization, and upstream section extraction remained important constraints.

## Introduction

Discharge summaries contain much of the information needed for clinical registries, pharmacoepidemiologic studies, and quality audits, but diagnoses, treatments, and investigation results are often recorded as free text. Clinical information extraction converts these narratives into structured data for secondary use. Previous studies have recovered stroke performance indicators from discharge summaries for registry audit[1, 2] and extracted features from clinical notes to supplement claims data in pharmacoepidemiology.[3, 4] Extending these uses to new health systems requires extraction methods that can accommodate local documentation practices and languages.

Kazakhstani discharge summaries illustrate this challenge. Our corpus contains 193,101 routine-care stroke summaries collected between 2013 and 2019, with dense Cyrillic text, abbreviations, list-like fragments, irregular formatting, and Russian–Kazakh mixing. Kazakh’s agglutinative morphology adds surface variation, as a single root can take many forms.[5, 6] These features compound the abbreviation and terminology problems documented in clinical text generally and Russian clinical text specifically.[7, 8]

The required output adds another layer of difficulty. Identifying a drug name is only the first step in assembling a medication record that links it to dosage, strength, form, route, frequency, and duration, sometimes across distant mentions.[9, 10] Similarly, a laboratory record must link the correct test, date, value, and unit despite variation in how results are written.[11] Locating document sections helps establish the context for these records and for related tasks such as comorbidity extraction.[12] We therefore distinguish detection, which identifies entities, from assembly, which links their fields into complete records. This distinction follows calls for clinical extraction benchmarks that better reflect downstream information needs.[13]

Evidence to guide these choices remains uneven. Clinical NLP research is concentrated in English and recurring institutional data sources, with named-entity recognition a common task.[14–16] Multilingual surveys and Russian benchmarks document progress in concept normalization, coding, and language understanding,[17, 18] while general-purpose Kazakh resources support language-model development.[5] Less is known about how these approaches compare when assembling records from mixed Russian–Kazakh clinical text. For institutions selecting an extraction system, this accuracy question is linked to practical choices about data residency, annotation effort, model adaptation, and computing resources.

Our team’s preliminary work approached this problem through clinical applications. Saduyeva et al. used an AI-based data-mining application with manual verification to obtain clinical variables from 152 HIV patient discharge records for mortality analysis.[19] Yermakov et al. used GPT-4o to extract medication and laboratory data from manually segmented sections of 272 stroke records, followed by manual review and correction, also for mortality analysis.[20]

These studies motivated a closer examination of the extraction process itself: how accurately different models identify and assemble records, and how performance changes when manually prepared sections are replaced by automatically constructed inputs.

Here, we compare a multilingual encoder, a locally fine-tuned small LLM, and a zero-shot frontier cloud model on section identification, medication extraction, and laboratory extraction using shared task-specific inputs and scoring. We assess detection and complete-record assembly separately, with annotator agreement providing an empirical reference for interpretation.[21] We then compare extraction from curated sections with a pipeline that starts from raw discharge summaries and examine its computational feasibility across the full corpus. This design connects model-level performance to the requirements of a retrospective clinical information system.

## Methods

### Study design, data, and cohorts

This offline retrospective evaluation used 193,101 anonymized Russian- and Kazakh-language stroke discharge summaries from one (Kazakhstani) health system (2013–2019). Extraction accuracy was assessed on existing task-specific annotated subsets. A separate corpus-wide operational analysis assessed processing coverage, parseability, and computational throughput.

Task 1 locates character-offset spans for lab_results, prescription, and drugs_hospital. The 4,327-document annotation collection has a group-aware 3,461/433/433 train/development/test split. Its predominantly all-three-label subset contains 2,482/320/332 documents; the 332-case consensus test set is the primary evaluation.

Task 2 extracts seven medication fields: drug name, dosage, strength, form, route, frequency, and duration. The source collection contains 708 human-verified examples. Its original test set has 154 example IDs but 152 canonical cases and 2,556 reference records after solvent exclusion. Exact ObjectId-to-case-ID mapping identified one case shared with training and two other cases each represented by multiple test examples with different texts and labels. All five affected examples were excluded by identity, identically across arms, leaving 149 unique cases and 2,475 reference records. This cohort is shared by primary benchmark and cascade analyses; the original cohort is retained as a sensitivity analysis.

Task 3 extracts (category, date, test, value, unit) quintuples. Its 1,888-document annotated collection is split into 1,507/190/191 training/validation/test documents, with all 50 double-annotated reports held out in the test set. Local models were trained on the corresponding training partition. All arms use this common 191-case test set and an output schema with 25 categories. All three tasks use internal data splits. Figure 1 summarizes the task cohorts, paired human samples, and corpus-scale processing population.

**Figure 1.**
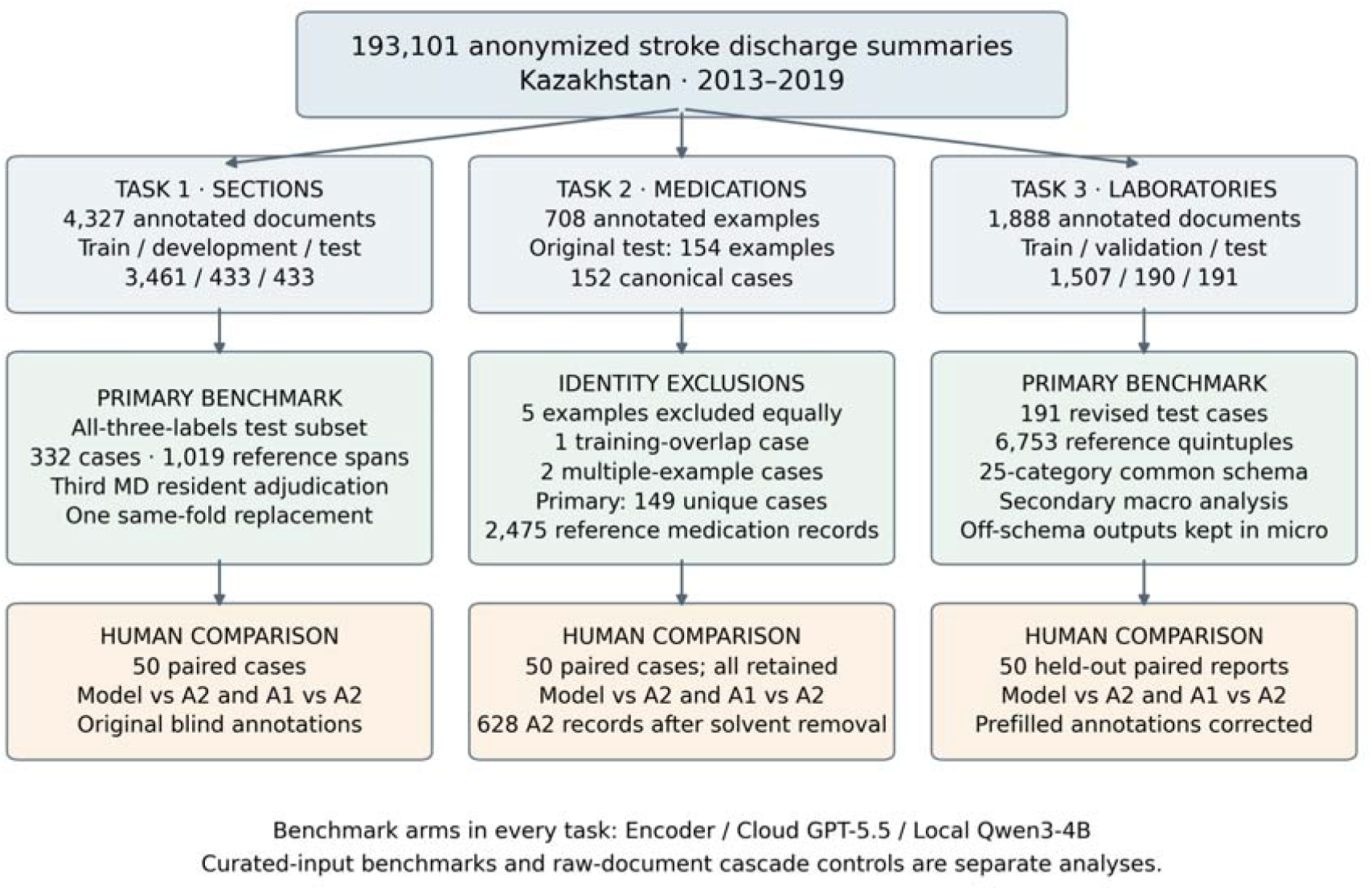
Study data flow and evaluation populations. Primary test cohorts contain 332 section, 149 medication, and 191 laboratory cases. Medication exclusions are identity-based and shared across models and cascade controls. Paired human comparisons use 50 held-out cases per task.

### Annotation and human reference

MD-resident annotators produced and checked the task annotations. Blind re-annotation used a block-selection interface for sections and empty record tables for medications and laboratories. A third MD resident assessed annotation disagreements to establish the final section reference labels. Human-agreement comparisons use the original annotations before adjudication: A1 is the original annotator and A2 the second annotator. Both A1 and each model are scored against A2 on the same documents.

Each task’s paired analysis includes 50 held-out cases; all 50 medication cases survive the primary exclusions. Common solvent filtering removes six of 634 A2 medication records, leaving 628 reference records.

### Task-specific representations and model families

For Task 1, a deterministic segmenter divides text into atomic blocks comprising lines, headers, list items, and sentence-split long lines. Models return labelled block-ID intervals (Figure 2), selecting spans from this shared representation.[22] For Tasks 2 and 3, generative models return structured records from medication or laboratory text. The benchmark supplies curated sections to evaluate record extraction, while the cascade constructs these inputs automatically from the full document.

**Figure 2.**
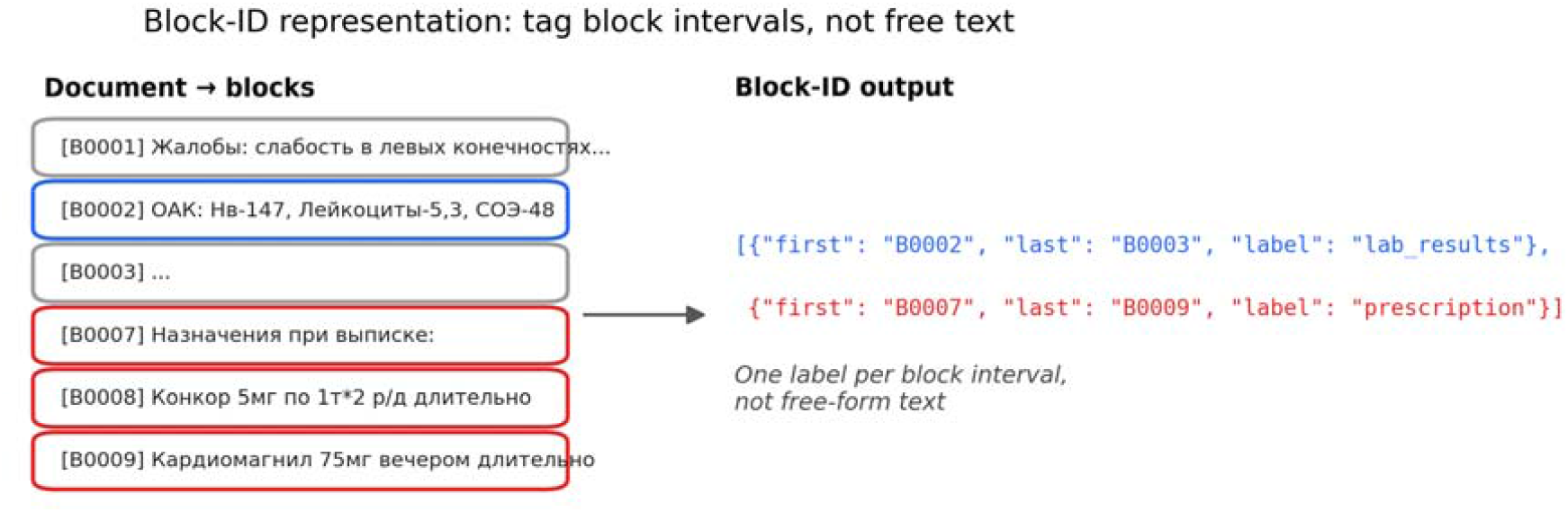
Task-1 block-ID representation. A document is split into atomic blocks, and the model identifies each target section by returning labelled intervals of block IDs.

#### Encoder

A multilingual ModernBERT-based encoder (mmBERT)[23] underwent masked-language-model domain-adaptive pretraining (DAPT) on approximately 100,000 unlabelled discharge reports from the same corpus. For sections, mean-pooled block embeddings feed a two-layer bidirectional LSTM (256 hidden units per direction) and a BIO linear-chain conditional random field (CRF). Training used eight epochs, batch size one with eight-step gradient accumulation, and validation macro-F1 checkpoint selection. The development programme included three seeds and CRF, presence-head, DAPT, and decoding ablations, reported separately in the supplementary material. Medication and laboratory encoder arms use field-specific sequence labelling and record post-processing.

#### Cloud model

GPT-5.5 (openai/gpt-5.5, accessed through OpenRouter; recorded evaluation period August 2026) was used zero-shot with section definitions or the relevant record schema. The section runner specifies temperature zero and a 2,048-token output limit. Laboratory extraction used temperature zero, JSON-object output, and a dictionary-based prompt augmented with annotation-guide rules for dates, categories, values, and units. Task-specific settings and prompts are documented in the supplement. Raw responses and per-document cost and latency were retained. Manual inspection of cloud-bound text for direct identifiers was performed. Identifier placeholders in the source text were used.

#### Local LLM

Per-task Qwen3-4B models[24] were adapted locally. Section training used four-bit NF4 QLoRA, rank 16, alpha 32, all-linear targets, three epochs, completion-only loss, and an 8,192-token context. Medication training used bf16 low-rank adaptation; laboratory training used four-bit QLoRA with rank 32, alpha 64, and five epochs. Both used validation-based checkpoint selection and greedy decoding. Recorded fine-tuning times ranged from approximately nine minutes to one hour on RTX 5090/H100 hardware.

### Outcomes and statistical analysis

Task-1 span detection uses same-label one-to-one bipartite matching at intersection-over-union ≥0.5, after human spans are snapped to the common blocks. Task-2 drug-name detection uses one-to-one normalized name matching, consuming duplicate names in record order, with the same solvent exclusion for both humans and every model.

The primary medication assembly outcome is normalized full-record recovery: matched records correct in all seven fields divided by all reference records, including unmatched references in the denominator. Field comparison uses parsed numeric/unit and frequency representations and resolved form/route codes, falling back to raw values when unresolved. Numeric comparison uses exact values without tolerance or cross-unit magnitude conversion. Drug names use trimmed, case-insensitive raw equality for assembly and normalized matching keys for detection. Dosage and strength are scored as separate fields. Raw-field recovery provides a sensitivity outcome with the same reference denominator.

Task-3 detection is micro-F1 over distinct normalized test names per document, irrespective of category. Assembly is micro-F1 over normalized quintuple sets. Unit canonicalization is identical for models and humans. Secondary macro-F1 averages over the same 25 declared categories for every arm, assigning zero when both gold and prediction support are empty. Primary micro scoring retains off-schema outputs. The supplementary material specifies the category keys and scoring rules.

Scores pool document-level sufficient counts before calculation. Marginal 95% percentile intervals use 2,000 document-bootstrap draws; paired differences use 10,000 shared document draws, seed 42. Two-sided centred-bootstrap tests use a plus-one finite-sample correction. Separate Holm families comprise three model–model or three model–human comparisons within each task/metric. Cascade families contain two medication-control contrasts or one laboratory-control contrast per metric. We report unadjusted percentile intervals alongside Holm-adjusted p-values.

### Raw-document system evaluation and reporting

The offline pipeline combines a consensus-v2 section encoder with laboratory keyword fallback, followed by per-task fine-tuned Qwen3-4B record extraction. Stage-1 offsets index the canonical full body text. The medication single-window variant selects a contiguous extracted window; the all-block variant concatenates extracted medication blocks. Matched curated controls use the same merged-model/vLLM stack and 3,400-token output budget as the medication cascade. This control differs from the separate adapter path used by the local benchmark arm.

The corpus-wide run measures processing coverage and computational demand through section-found rates, parseability conditional on finding a section, record counts, and recorded GPU-hours. Accuracy is assessed separately on the labelled cascade cohort.

A supplementary reporting map covers applicable TRIPOD-LLM domains[25] and journal-specific provenance and reproducibility requirements. The supplement also documents experimental settings and prompts. AI assistance with evaluation-code development and manuscript copyediting is disclosed in Statements and Declarations.

## Results

### Detection and record assembly

Table 1 reports primary pooled estimates. Section-span F1 is 0.919 for the encoder, 0.926 for the cloud model, and 0.932 for the local LLM, over 1,019 reference spans. The local-minus-cloud difference is 0.006 (95% CI −0.010 to 0.021; Holm p=0.804), with the interval spanning differences in either direction.

**Table 1.** Primary test-set performance, estimate [95% document-bootstrap CI]. Columns share cases and metric definitions within each row. Medication recovery uses all 2,475 reference records; laboratory macro-F1 is secondary and schema-restricted.

| Metric (cases) | Encoder | Cloud GPT-5.5 | Local Qwen3-4B |
| --- | --- | --- | --- |
| Section-span F1 (332) | 0.919 [0.900, 0.936] | 0.926 [0.908, 0.942] | **0.932** [0.915, 0.947] |
| Medication name F1 (149) | 0.917 [0.904, 0.930] | **0.966** [0.956, 0.975] | 0.940 [0.914, 0.960] |
| Medication normalized recovery (149) | 0.095 [0.066, 0.126] | 0.311 [0.266, 0.358] | **0.381** [0.328, 0.436] |
| Laboratory test-name F1 (191) | 0.851 [0.841, 0.861] | 0.960 [0.954, 0.966] | **0.966** [0.960, 0.973] |
| Laboratory quintuple F1 (191) | 0.474 [0.431, 0.515] | 0.822 [0.795, 0.847] | **0.892** [0.874, 0.909] |
| Laboratory schema macro-F1 (191) | 0.112 [0.101, 0.121] | 0.404 [0.327, 0.433] | **0.425** [0.364, 0.443] |

Medication model rankings change with the output requirement. The cloud model leads drug-name detection, with a local-minus-cloud difference of −0.026 [−0.046, −0.010] (Holm p=0.011). Complete normalized recovery instead favours the local model: 0.381 versus 0.311, difference +0.070 [0.026, 0.114] (Holm p=0.002). Raw-field recovery is 0.080/0.173/0.284 for encoder/cloud/local, showing sensitivity to normalization on the same cohort. In the original 154-example cohort, before identity-based exclusions, normalized recovery is 0.092/0.310/0.369.

For laboratories, local-minus-cloud test-name F1 is +0.006 [0.000, 0.012], whereas the quintuple difference is +0.070 [0.047, 0.094]. The encoder scored lower than both generative arms on detection and assembly. On the common 25-category schema, local-minus-cloud macro-F1 is +0.021 [−0.025, 0.073]. The category-support analysis provides no clear overall separation of the generative arms on this secondary endpoint. Encoder/cloud/local outputs contain 565/zero/20 off-schema quintuples, retained in primary micro scoring. Supplementary tables and Figure S1 report category support, full pairwise contrasts, and sensitivity analyses.

### Paired comparison with annotator agreement

Human agreement is 0.905 for section-span F1 and 0.959 for medication detection on the respective 50-case samples. Normalized medication recovery for A1 against A2 is 0.178. Table 2 and Figure 3 report model-minus-human differences using the same reference and cases.

**Table 2.** Paired model-minus-human differences [95% CI], followed by Holm-adjusted p-values. The estimand is (model versus A2) minus (A1 versus A2), using 50 held-out cases per task.

| Metric (paired cases) | Encoder | Cloud GPT-5.5 | Local Qwen3-4B |
| --- | --- | --- | --- |
| Section-span F1 (50) | +0.006 [−0.006, +0.019];<br>.421 | −0.023 [−0.060, +0.011];<br>.421 | +0.018 [+0.003, +0.041];<br>.204 |
| Medication name F1 (50) | −0.030 [−0.050, −0.008];<br>.018 | +0.020 [+0.006, +0.038];<br>.029 | −0.005 [−0.034, +0.018];<br>.676 |
| Medication normalized recovery (50) | −0.084 [−0.138, −0.032];<br>.002 | +0.175 [+0.080, +0.274];<br><.001 | +0.172 [+0.082, +0.264];<br><.001 |
| Laboratory test-name F1 (50) | −0.118 [−0.136, −0.103];<br><.001 | +0.006 [−0.001, +0.013];<br>.097 | −0.009 [−0.016, −0.002];<br>.039 |
| Laboratory quintuple F1 (50) | −0.440 [−0.502, −0.381];<br><.001 | −0.069 [−0.121, −0.029];<br>.015 | −0.003 [−0.020, +0.013];<br>.700 |

The generative medication arms agree more closely with A2 on normalized complete records than A1 does. On the 50 laboratory reports, human quintuple agreement is 0.902 and local-model agreement with A2 is 0.898, with a paired difference of −0.003 [−0.020, +0.013].

**Figure 3.**
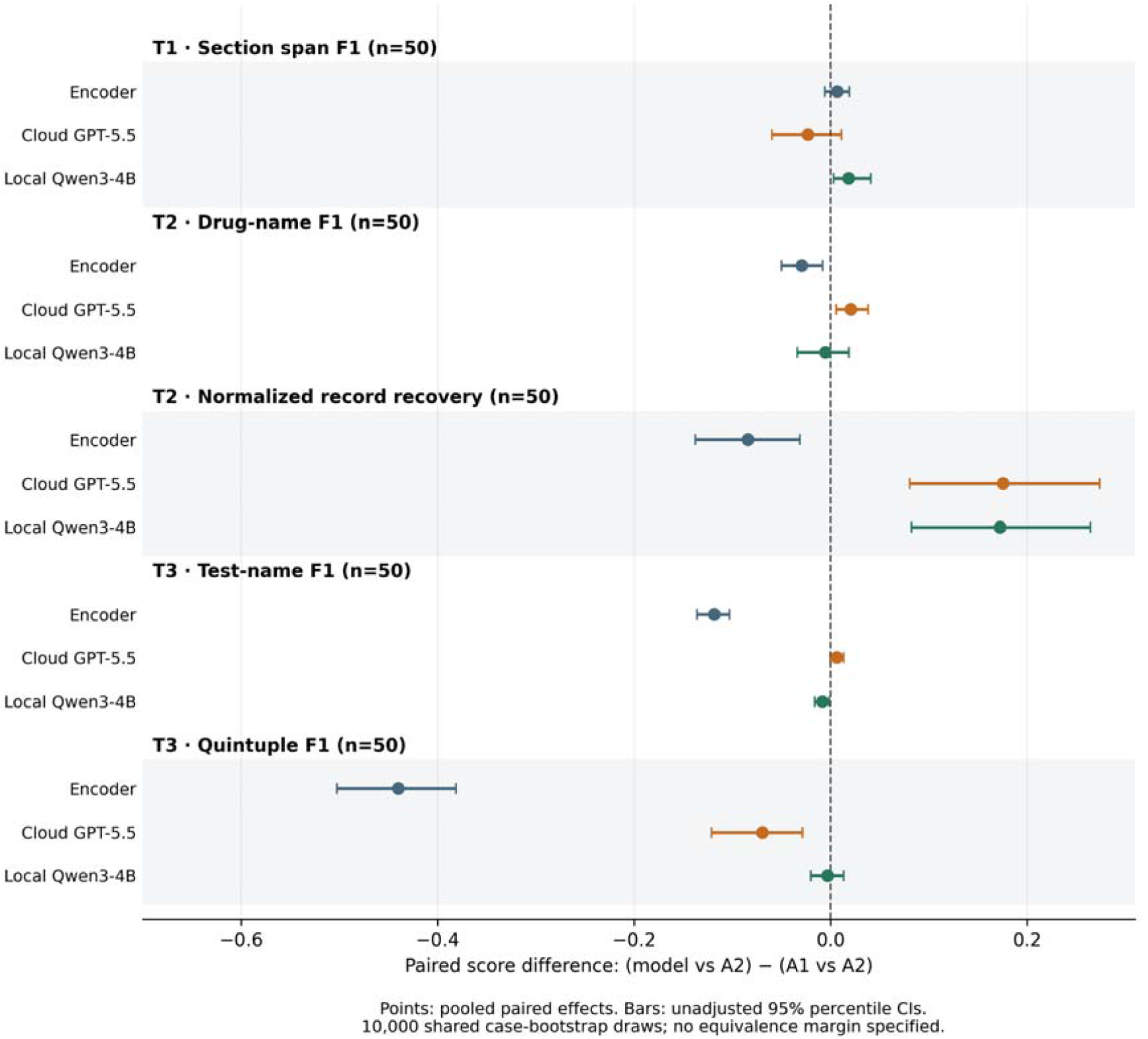
Paired model-minus-human differences using A2 as the common reference on 50 held-out cases per task. Points and intervals are pooled differences and unadjusted 95% paired document-bootstrap intervals. Zero denotes equal observed agreement. Table 2 supplies Holm-adjusted p-values.

### Raw-document cascade and corpus processing

Automatic section extraction reduces record-level performance (Table 3). Relative to matched curated controls, medication recovery changes by −0.173 [−0.216, −0.131] for one window and −0.131 [−0.180, −0.084] for all blocks. Laboratory quintuple F1 changes by −0.087 [−0.111, −0.065]. Saved error inspection also found drug-name omissions when the name remained in the supplied section, distinguishing record-generation failures from upstream content loss.

**Table 3.** Matched curated controls and raw-document cascades. Medication assembly is normalized full-record recovery; laboratory assembly is quintuple F1. Medication controls and cascades use the same merged-model/vLLM stack, distinct from the benchmark adapter path in Table 1. The laboratory comparison retains its original model and 191-case cohort, which shares 149 cases with the Table 1 benchmark.

| Task and input | Cases | Name F1 | Assembly |
| --- | --- | --- | --- |
| Medication curated control | 149 | 0.921 | 0.377 |
| Medication single-window cascade | 149 | 0.752 | 0.204 |
| Medication all-block cascade | 149 | 0.779 | 0.246 |
| Laboratory curated control | 191 | 0.965 | 0.898 |
| Laboratory raw-document cascade | 191 | 0.917 | 0.811 |

After excluding 116 partition IDs absent from the canonical population, the corpus-wide analysis covered all 193,101 cases. The original pipeline produced 2,540,326 medication records and 5,271,287 laboratory tuples in 13.3 H100 GPU-hours. Table 4 reports section coverage and the proportion of section-found outputs that were parseable. Recorded cloud benchmark charges were approximately US$0.080 per medication input and US$0.085 per laboratory input. These charges apply to curated-section extraction, whereas the local runtime covers the full pipeline.

**Table 4.**
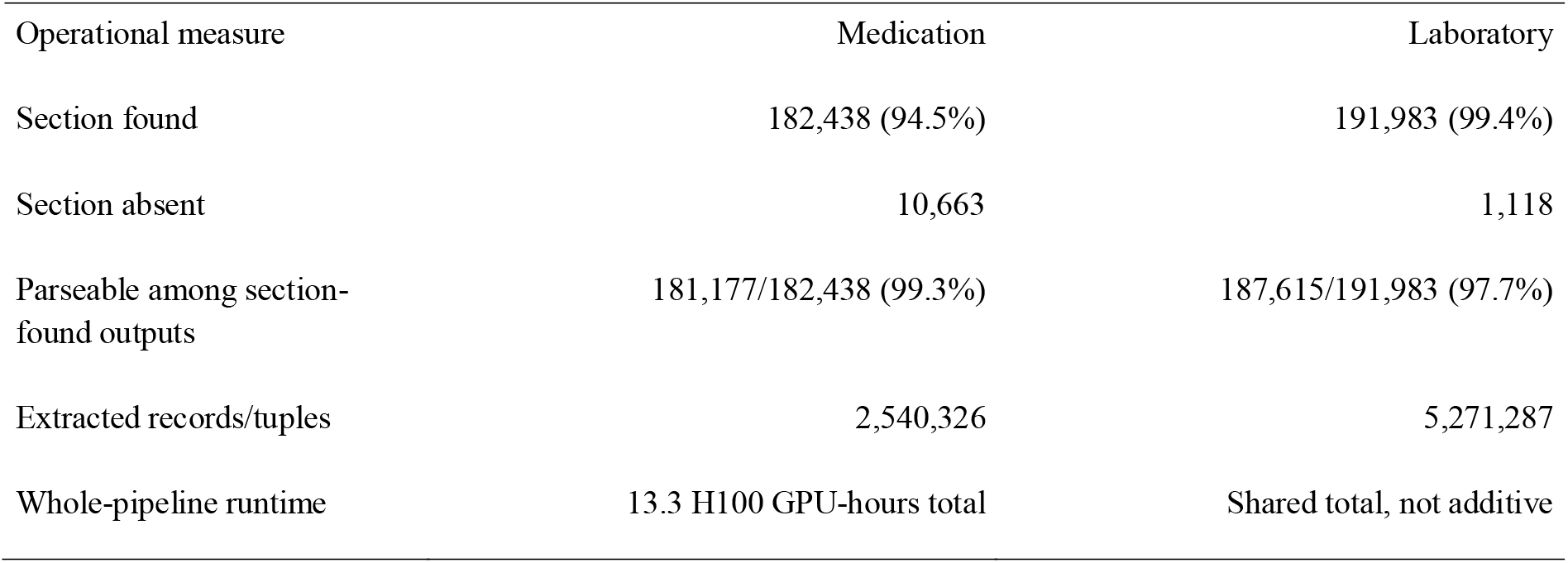
Canonical corpus-scale operational results (193,101 summaries). Parseability is calculated among section-found outputs. Runtime is the recorded total for the original local pipeline.

| Operational measure | Medication | Laboratory |
| --- | --- | --- |
| Section found | 182,438 (94.5%) | 191,983 (99.4%) |
| Section absent | 10,663 | 1,118 |
| Parseable among section-found outputs | 181,177/182,438 (99.3%) | 187,615/191,983 (97.7%) |
| Extracted records/tuples | 2,540,326 | 5,271,287 |
| Whole-pipeline runtime | 13.3 H100 GPU-hours total | Shared total, not additive |

## Discussion

### Implications for clinical information systems

The preferred extraction model depended on how much detail the output needed to preserve. The compact encoder was competitive for locating clinical sections, whereas the fine-tuned local LLM performed best on complete medication and laboratory records. The cloud model achieved the highest medication-name detection score. Together, these findings show how a model that is well suited to identifying mentions may differ from one that is best suited to linking their attributes. They extend reports of clinical extraction with general and local LLMs[26–28] by comparing these output requirements within the same clinical corpus.

This distinction matters when choosing a system for secondary use. An application that indexes sections or identifies candidate mentions places different demands on extraction from one that studies medication doses or follows laboratory values over time. In our comparisons, these more detailed outputs favoured local adaptation. Local inference also keeps processing within institutional infrastructure, although it requires labelled examples, hardware, and maintenance. Cloud inference offers an alternative when adaptation resources are limited and external processing is permitted. The relevant choice therefore combines the required record detail with the resources and processing arrangements available to the institution.

Performance also depended on the text supplied to the extraction model. Starting from raw documents introduced losses when sections were found, joined, or presented to the downstream model. Retaining all medication blocks recovered some of the performance lost with a single window, but remained below the matched curated-input control. Drug-name omissions also occurred when the name was still present in the supplied text, indicating a combination of upstream content loss and downstream extraction errors. Component benchmarks and raw-document evaluation thus provide complementary information: the former measures extraction from prepared inputs, while the latter captures the additional difficulties introduced by the processing pipeline.

### Annotation conventions and record fidelity

Even with curated inputs, assembling medication records remained difficult. The best normalized recovery was 0.381, meaning that fewer than two in five reference records matched across all seven fields. Raw-field recovery was lower, at 0.284. This gap between identifying a drug and recovering its full record is consistent with earlier medication-extraction research.[9, 10] Because the assembly criterion requires every field to agree, a discrepancy in a single attribute is enough for an otherwise well-matched record to fail.

The qualitative review helps explain these discrepancies. Models and annotators differed in assigning quantities to dosage or strength, recording inferred or verbatim forms and routes, expressing frequency, and delimiting laboratory sections. Normalization reconciled some surface differences while preserving distinctions between fields. Reporting both raw and normalized recovery therefore shows how the scoring rules affect measured performance. It also helps relate benchmark results to the information a downstream application needs, a recurring concern in clinical benchmark design.[29]

Annotator agreement provides a complementary view of this uncertainty. Higher agreement with the reference annotations than that achieved by an individual annotator may reflect more consistent application of the coding scheme.[21] The qualitative overlap between annotation disagreements and model errors suggests that some extraction difficulties are shared. In practice, reviewing complete records requires attention to both the extracted content and the interpretation encoded in the reference labels.

## Limitations

Several features of the evaluation limit its generalizability. All splits were drawn internally from one disease domain and health system, so performance on external institutions or later documentation remains untested. The 2013–2019 corpus was evaluated using 2026-era models. The section test cohort predominantly contained all target labels, providing limited evidence about documents with missing sections. Human comparisons used 50 reports per task. Language-specific extraction performance also remains unestablished because the available character-based language identification provides only a coarse description of the text.

Data exposure and differences between modelling approaches also affect interpretation. Identity checks removed known case overlap and ambiguous repeated examples, but incomplete patient-level linkage leaves possible overlap between records from the same patient. DAPT included test text without labels; the non-random Task-1 exposed and unexposed groups contained 179 and 153 cases, respectively, with encoder F1 rounding to 0.919 in both. This comparison cannot separate an exposure effect from differences between the groups. Similarly, comparing a selected fine-tuning recipe with a zero-shot cloud prompt estimates the performance of those approaches as used. The separate contributions of model family, capacity, training exposure, prompting, annotation ambiguity, and normalization remain unresolved.

The evaluation also has limits for downstream use. Category aliases and empty-support categories complicate interpretation of laboratory macro-F1, and agreement with reference records does not directly measure clinical usability. The runtime and cloud charges cover different hardware and processing scopes and exclude governance and human-review costs, limiting cost comparisons. Finally, the study assessed offline extraction and throughput; clinical workflow effectiveness, safety, and patient benefit require separate evaluation.

## Conclusion

In this Russian–Kazakh stroke-discharge benchmark, local models performed strongly on detection-focused tasks, and fine-tuned local LLMs achieved the best complete-record assembly scores. The cloud model led medication-name detection. These task-dependent results support choosing an extraction approach according to the required record detail and institutional processing constraints. Performance losses between curated sections and raw documents further show the importance of evaluating the full extraction pathway. Reliable secondary use depends on both identifying the relevant information and preserving its relationships in the resulting records.

## Data Availability

Annotated data are available under institutional data-use agreement; requests should be directed to the corresponding author.

## Statements and Declarations

### Ethics approval

The Institutional Review Ethics Committee of Nazarbayev University approved this retrospective study (NU-IREC 833/10012024; January 22, 2024).

### Consent to participate

The ethics committee waived informed consent for the retrospective study.

### Consent for publication

The manuscript reports aggregate results and non-identifying examples.

### Data availability

The anonymized corpus may be made available under an institutional data-use agreement upon request to the study principal investigator through the corresponding author. Access is subject to institutional approval.

### Material and code availability

An evaluation-code supplement includes scoring/statistical source, normalization dictionaries, and a runnable synthetic-count example. It contains no clinical documents, annotations, predictions, or trained weights. Empirical reproduction requires authorized data access. Supplementary material provides detailed evaluation definitions, sensitivity results, prompts/settings recoverable from study records, and a reporting map.

### Author contributions

AT: Conceptualization, Data curation, Formal analysis, Investigation, Methodology, Software, Writing - Original Draft; IA: Data curation, Formal analysis, Investigation, Methodology, Software, Writing - Original Draft; GZ: Investigation, Validation, Visualization, Writing - Review & Editing; AR: Validation, Visualization, Writing - Review & Editing; SS: Data curation, Validation, Writing - Review & Editing; AG: Conceptualization, Funding acquisition, Project administration, Resources, Supervision, Writing - Review & Editing;

### Use of artificial intelligence

AI assistance with analysis code and manuscript copyediting are disclosed in Methods. GLM models (5.1-5.3) were used for code assistance, ChatGPT was used for copyediting. Human authors retain responsibility for the analyses and final text.

