## Supplementary material for "Task-dependent model selection for structured extraction from multilingual non-English clinical records": supplementary_material_jmedsys_5.docx

### S1. Evaluation population, targets, and reference identity

This supplement describes the evaluation populations, scoring procedures, sensitivity analyses, and experimental settings for the retrospective extraction study. The primary benchmark compares Encoder, Cloud GPT-5.5, and Local Qwen3-4B on 332 section-extraction cases, 149 medication cases, and 191 laboratory cases.

The source population contains 193,101 anonymized stroke discharge summaries from Kazakhstani hospitals, dated 2013–2019. Each task uses its own annotated subset of this population. Task 1 contains 4,327 annotated documents, with 3,461/433/433 train/development/test documents. Its three-label subset contains 2,482/320/332 documents and provides limited coverage of documents with absent sections. The three section labels are lab_results, prescription, and drugs_hospital. Task 2 contains 708 annotated examples; its original test export contains 154 examples. Task 3 contains 1,888 annotated documents, with a 1,507/190/191 train/validation/test split.

#### Medication case-identity reconciliation

Medication export identifiers use more than one identifier namespace. Exact mapping to canonical cases identifies 152 cases among 154 test examples. One case also occurs in training; two other cases each have multiple test examples with different texts and labels. All five affected examples are excluded by identity alone, equally for every arm. The resulting primary population contains 149 unique cases and 2,475 reference medication records after the common solvent rule. All 50 medication human-comparison cases are retained. S4 reports a sensitivity analysis on the original 154-example population, which contains 2,556 reference records. Medication cascade-control comparisons use the same 149-case primary population.

Task 1 used group-aware splitting to address identified duplicates and shared-patient/source groups. Medication case-identity exclusions similarly remove known overlaps and repeated examples. Incomplete patient-level linkage leaves possible overlap between longitudinal records from the same patient.

#### Annotation references

For Task 1, a third MD resident assessed annotation disagreements to establish the final section reference labels. One out-of-scope EMR export was replaced with a discharge summary from the same test fold, with replacement-case predictions supplied for all arms. The scored reference contains 1,019 spans. Human-agreement comparisons use the original annotations before adjudication.

A1 and A2 denote designated protocol roles: A1 is the original reference annotation and A2 the second annotation. A2 was obtained by blind re-annotation for all tasks. Both A1 and each model are scored against the identical A2 annotations on identical cases. Each task has 50 paired cases. All 50 Task 3 double-annotated reports are held out in the 191-case model test set. The A2 medication export contains 634 records before the common solvent exclusion and 628 afterward.

### S2. Metric definitions and statistical analysis

#### Task-specific scoring

**Task 1: section localization.** Same-label reference and predicted spans are matched one-to-one by bipartite matching, with intersection-over-union at least 0.5. Human spans are snapped to the same block representation used by the models. True-positive, false-positive, and false-negative counts are pooled over documents before computing micro-F1.

**Task 2: medication-name detection.** The official scorer performs one-to-one normalized drug-name matching, consuming duplicate names in record order. Name normalization uses drug-dictionary aliases, Latin-prefix handling, substring matching, and guarded fuzzy matching, with a trimmed lower-case raw-name fallback. The same deterministic solvent filter is applied to A1, A2, and every model before scoring. Detection is pooled micro-F1 and includes unmatched predictions as false positives and unmatched reference records as false negatives.

**Task 2: normalized full-record recovery.** Seven fields are required: drug name, dosage, strength, form, route, frequency, and duration. The numerator is the number of matched records for which all seven fields agree; the denominator is *all reference records*, including unmatched references. False-positive records affect detection F1 but do not directly enter the recovery calculation. Reporting detection and recovery together captures both extractions unsupported by the reference and incomplete recovery of reference records.

For dosage, strength, and duration, exact parsed numeric-value/unit-code tuples are compared when a numeric value is available; otherwise trimmed case-insensitive raw values are used. There is no numeric tolerance or cross-unit magnitude conversion in this comparison. Frequency uses the parsed (times_per_dose, doses_per_day, timing) tuple when any component is available, otherwise the raw value. Form and route use resolved vocabulary codes, falling back to raw values. Drug-name equality for assembly remains trimmed and case-insensitive *raw-name* equality, even though detection uses a normalized matching key. Dosage and strength remain separate slots; an equivalent quantity in the wrong slot fails. Null, empty, textual null/None, hyphen, and scalar NaN representations are treated as missing under the official field parser; two missing values compare equal.

**Task 2: raw-field sensitivity.** The same name matches and all-reference denominator are retained, but all seven fields use trimmed case-insensitive raw comparison with the same missing-value treatment. Comparing raw and normalized recovery shows the effect of reconciling field representations during scoring.

**Task 3: laboratory detection and assembly.** Detection compares sets of distinct normalized test names within each document, ignoring category, date, value, and unit. Assembly compares normalized (category, date, test, value, unit) quintuple sets. Repeated identical items within a document are deduplicated by the set representation; counts are then pooled across documents. The same historical unit-canonicalization prepass and official normalized tuple conversion are used for all models and both annotators. Primary micro scores include off-schema outputs. For name detection, an off-schema output can still match a gold test name because category is explicitly not part of this endpoint; for quintuple assembly its category must agree.

**Task 3: common-schema macro sensitivity.** The declared output schema contains 25 literal categories, including rh_facto, and is held fixed across arms and every bootstrap resample. Each category contributes its pooled quintuple F1 with equal weight; if both gold and predicted support are zero, its F1 is zero. The schema-restricted macro analysis excludes off-schema categories, while primary micro scoring retains them. Category keys are matched literally, so rh_facto and rh_factor remain distinct. Macro-F1 is a secondary endpoint.

#### Aggregation, intervals, and comparisons

For F1, the pooled score is $2TP/(2TP+FP+FN)$. Medication recovery is $C/G$, where $C$ denotes all-seven-field-correct records and $G$ denotes all reference records. Document-level sufficient counts, rather than means of individual document scores, are resampled. Marginal 95% percentile confidence intervals use 2,000 document-bootstrap draws. Paired differences use 10,000 shared document draws and seed 42, recalculating both pooled scores on each shared draw. Human contrasts are (model vs A2) − (A1 vs A2); model contrasts are the named first arm minus the second arm on the corresponding benchmark reference.

Two-sided bootstrap p-values use a null-centred distribution and a plus-one finite-sample correction. Holm adjustment is performed separately for each task/metric and comparison type: three model–model contrasts and three model–human contrasts form separate families. The two medication cascade-control contrasts form one family per metric; the single laboratory cascade-control contrast forms a one-comparison family per metric. Confidence intervals are unadjusted percentile intervals, whereas p-values incorporate multiplicity adjustment. This can yield an interval excluding zero alongside an adjusted p-value above 0.05. P-values below 0.001 are displayed as <0.001. All comparisons use difference tests; equivalence and non-inferiority margins were not specified.

### S3. Complete benchmark and paired comparisons

**Table S1. Marginal pooled benchmark estimates and 95% confidence intervals.**

| Metric | Arm | Cases | Estimate [95% CI] | TP/FP/FN or correct/reference |
| --- | --- | --- | --- | --- |
| T1 span F1 | Encoder | 332 | 0.919 [0.900, 0.936] | 917/60/102 |
| T1 span F1 | Cloud GPT-5.5 | 332 | 0.926 [0.908, 0.942] | 953/86/66 |
| T1 span F1 | Local Qwen3-4B | 332 | 0.932 [0.915, 0.947] | 927/44/92 |
| T2 drug-name F1 | Encoder | 149 | 0.917 [0.904, 0.930] | 2285/223/190 |
| T2 drug-name F1 | Cloud GPT-5.5 | 149 | 0.966 [0.956, 0.975] | 2452/149/23 |
| T2 drug-name F1 | Local Qwen3-4B | 149 | 0.940 [0.914, 0.960] | 2336/160/139 |
| T2 normalized record recovery | Encoder | 149 | 0.095 [0.066, 0.126] | 234/2475 |
| T2 normalized record recovery | Cloud GPT-5.5 | 149 | 0.311 [0.266, 0.358] | 769/2475 |
| T2 normalized record recovery | Local Qwen3-4B | 149 | 0.381 [0.328, 0.436] | 942/2475 |
| T3 test-name F1 | Encoder | 191 | 0.851 [0.841, 0.861] | 4305/667/840 |
| T3 test-name F1 | Cloud GPT-5.5 | 191 | 0.960 [0.954, 0.966] | 4925/187/220 |
| T3 test-name F1 | Local Qwen3-4B | 191 | 0.966 [0.960, 0.973] | 4973/173/172 |
| T3 quintuple F1 | Encoder | 191 | 0.474 [0.431, 0.515] | 3189/3507/3564 |
| T3 quintuple F1 | Cloud GPT-5.5 | 191 | 0.822 [0.795, 0.847] | 5546/1190/1207 |
| T3 quintuple F1 | Local Qwen3-4B | 191 | 0.892 [0.874, 0.909] | 6024/732/729 |
| T3 schema macro-F1 (secondary) | Encoder | 191 | 0.112 [0.101, 0.121] | 25 categories |
| T3 schema macro-F1 (secondary) | Cloud GPT-5.5 | 191 | 0.404 [0.327, 0.433] | 25 categories |
| T3 schema macro-F1 (secondary) | Local Qwen3-4B | 191 | 0.425 [0.364, 0.443] | 25 categories |

Counts refer to the metric-specific representation. Task 3 has 5,145 distinct test-name references pooled over documents and 6,753 reference quintuples.

**Table S2. Complete paired model–model contrasts.**

| Metric | First minus second arm | Difference [95% CI] | Holm p |
| --- | --- | --- | --- |
| T1 span F1 | Cloud-Encoder | +0.007 [-0.010, +0.025] | 0.804 |
| T1 span F1 | Local-Encoder | +0.013 [-0.002, +0.028] | 0.268 |
| T1 span F1 | Local-Cloud | +0.006 [-0.010, +0.021] | 0.804 |
| T2 drug-name F1 | Cloud-Encoder | +0.049 [+0.037, +0.061] | <0.001 |
| T2 drug-name F1 | Local-Encoder | +0.023 [-0.003, +0.045] | 0.062 |
| T2 drug-name F1 | Local-Cloud | -0.026 [-0.046, -0.010] | 0.011 |
| T2 normalized record recovery | Cloud-Encoder | +0.216 [+0.181, +0.253] | <0.001 |
| T2 normalized record recovery | Local-Encoder | +0.286 [+0.238, +0.335] | <0.001 |
| T2 normalized record recovery | Local-Cloud | +0.070 [+0.026, +0.114] | 0.002 |
| T3 test-name F1 | Cloud-Encoder | +0.109 [+0.100, +0.119] | <0.001 |
| T3 test-name F1 | Local-Encoder | +0.115 [+0.107, +0.124] | <0.001 |
| T3 test-name F1 | Local-Cloud | +0.006 [+0.000, +0.012] | 0.034 |
| T3 quintuple F1 | Cloud-Encoder | +0.348 [+0.313, +0.385] | <0.001 |
| T3 quintuple F1 | Local-Encoder | +0.418 [+0.381, +0.457] | <0.001 |
| T3 quintuple F1 | Local-Cloud | +0.070 [+0.047, +0.094] | <0.001 |
| T3 schema macro-F1 (secondary) | Cloud-Encoder | +0.292 [+0.216, +0.323] | <0.001 |
| T3 schema macro-F1 (secondary) | Local-Encoder | +0.313 [+0.251, +0.334] | <0.001 |
| T3 schema macro-F1 (secondary) | Local-Cloud | +0.021 [-0.025, +0.073] | 0.405 |

The local-minus-cloud medication-name effect favours the cloud arm, whereas the medication-recovery and laboratory-quintuple effects favour the local arm. The confidence interval for the common-schema macro difference spans effects in either direction.

**Table S3. Marginal scores on paired cases, all against A2.**

| Metric | Paired cases | Scored annotation/model | Estimate [95% CI] |
| --- | --- | --- | --- |
| T1 span F1 | 50 | A1 | 0.905 [0.838, 0.967] |
| T1 span F1 | 50 | Encoder | 0.911 [0.848, 0.971] |
| T1 span F1 | 50 | Cloud GPT-5.5 | 0.882 [0.821, 0.939] |
| T1 span F1 | 50 | Local Qwen3-4B | 0.923 [0.866, 0.976] |
| T2 drug-name F1 | 50 | A1 | 0.959 [0.940, 0.975] |
| T2 drug-name F1 | 50 | Encoder | 0.929 [0.908, 0.948] |
| T2 drug-name F1 | 50 | Cloud GPT-5.5 | 0.979 [0.967, 0.989] |
| T2 drug-name F1 | 50 | Local Qwen3-4B | 0.953 [0.924, 0.975] |
| T2 normalized record recovery | 50 | A1 | 0.178 [0.117, 0.253] |
| T2 normalized record recovery | 50 | Encoder | 0.094 [0.046, 0.159] |
| T2 normalized record recovery | 50 | Cloud GPT-5.5 | 0.354 [0.263, 0.446] |
| T2 normalized record recovery | 50 | Local Qwen3-4B | 0.350 [0.263, 0.437] |
| T3 test-name F1 | 50 | A1 | 0.976 [0.968, 0.985] |
| T3 test-name F1 | 50 | Encoder | 0.858 [0.840, 0.873] |
| T3 test-name F1 | 50 | Cloud GPT-5.5 | 0.982 [0.974, 0.989] |
| T3 test-name F1 | 50 | Local Qwen3-4B | 0.968 [0.957, 0.978] |
| T3 quintuple F1 | 50 | A1 | 0.902 [0.883, 0.921] |
| T3 quintuple F1 | 50 | Encoder | 0.461 [0.395, 0.523] |
| T3 quintuple F1 | 50 | Cloud GPT-5.5 | 0.832 [0.781, 0.874] |
| T3 quintuple F1 | 50 | Local Qwen3-4B | 0.898 [0.878, 0.918] |

On the 50 held-out Task 3 double-annotated reports, A1-versus-A2 agreement is test-name F1 0.976 [0.968, 0.985] and quintuple F1 0.902 [0.883, 0.921]. These estimates provide the human baseline for the model-minus-human contrasts on the same reports.

**Table S4. Complete paired model-minus-human effects with common A2 reference.**

| Metric | Paired cases | Arm | Difference [95% CI] | Holm p |
| --- | --- | --- | --- | --- |
| T1 span F1 | 50 | Encoder | +0.006 [-0.006, +0.019] | 0.421 |
| T1 span F1 | 50 | Cloud GPT-5.5 | -0.023 [-0.060, +0.011] | 0.421 |
| T1 span F1 | 50 | Local Qwen3-4B | +0.018 [+0.003, +0.041] | 0.204 |
| T2 drug-name F1 | 50 | Encoder | -0.030 [-0.050, -0.008] | 0.018 |
| T2 drug-name F1 | 50 | Cloud GPT-5.5 | +0.020 [+0.006, +0.038] | 0.029 |
| T2 drug-name F1 | 50 | Local Qwen3-4B | -0.005 [-0.034, +0.018] | 0.676 |
| T2 normalized record recovery | 50 | Encoder | -0.084 [-0.138, -0.032] | 0.002 |
| T2 normalized record recovery | 50 | Cloud GPT-5.5 | +0.175 [+0.080, +0.274] | <0.001 |
| T2 normalized record recovery | 50 | Local Qwen3-4B | +0.172 [+0.082, +0.264] | <0.001 |
| T3 test-name F1 | 50 | Encoder | -0.118 [-0.136, -0.103] | <0.001 |
| T3 test-name F1 | 50 | Cloud GPT-5.5 | +0.006 [-0.001, +0.013] | 0.097 |
| T3 test-name F1 | 50 | Local Qwen3-4B | -0.009 [-0.016, -0.002] | 0.039 |
| T3 quintuple F1 | 50 | Encoder | -0.440 [-0.502, -0.381] | <0.001 |
| T3 quintuple F1 | 50 | Cloud GPT-5.5 | -0.069 [-0.121, -0.029] | 0.015 |
| T3 quintuple F1 | 50 | Local Qwen3-4B | -0.003 [-0.020, +0.013] | 0.700 |

For Task 3 quintuple F1, the encoder and cloud model score below the A1-versus-A2 baseline; the local-minus-human interval includes differences in either direction. Figure 3 in the main article displays these paired differences. Human agreement measures annotation reproducibility under the study protocol.

### S4. Medication normalization and original-cohort sensitivity

Table S5 separates the effects of cohort selection and field normalization. All rows use the same deterministic solvent exclusion. The original 154-example cohort includes the overlapping and repeated cases described in S1, while the primary cohort excludes them. Within the primary cohort, raw and normalized recovery quantify the effect of field-scoring conventions.

**Table S5. Medication scoring and population sensitivities.**

| Arm | Population | Reference records | Metric | Estimate [95% CI] |
| --- | --- | --- | --- | --- |
| Encoder | Primary 149 | 2475 | Drug-name F1 | 0.917 [0.904, 0.930] |
| Encoder | Primary 149 | 2475 | Raw recovery | 0.080 [0.053, 0.111] |
| Encoder | Primary 149 | 2475 | Normalized recovery | 0.095 [0.066, 0.126] |
| Encoder | Original 154 | 2556 | Drug-name F1 | 0.918 [0.905, 0.930] |
| Encoder | Original 154 | 2556 | Normalized recovery | 0.092 [0.064, 0.124] |
| Cloud GPT-5.5 | Primary 149 | 2475 | Drug-name F1 | 0.966 [0.956, 0.975] |
| Cloud GPT-5.5 | Primary 149 | 2475 | Raw recovery | 0.173 [0.133, 0.215] |
| Cloud GPT-5.5 | Primary 149 | 2475 | Normalized recovery | 0.311 [0.266, 0.358] |
| Cloud GPT-5.5 | Original 154 | 2556 | Drug-name F1 | 0.966 [0.957, 0.975] |
| Cloud GPT-5.5 | Original 154 | 2556 | Normalized recovery | 0.310 [0.265, 0.358] |
| Local Qwen3-4B | Primary 149 | 2475 | Drug-name F1 | 0.940 [0.914, 0.960] |
| Local Qwen3-4B | Primary 149 | 2475 | Raw recovery | 0.284 [0.236, 0.335] |
| Local Qwen3-4B | Primary 149 | 2475 | Normalized recovery | 0.381 [0.328, 0.436] |
| Local Qwen3-4B | Original 154 | 2556 | Drug-name F1 | 0.932 [0.906, 0.954] |
| Local Qwen3-4B | Original 154 | 2556 | Normalized recovery | 0.369 [0.316, 0.422] |

For the 50-case same-A2 comparison, raw recovery is 0.132 [0.074, 0.204] for A1, 0.088 [0.039, 0.152] for Encoder, 0.299 [0.212, 0.391] for Cloud GPT-5.5, and 0.306 [0.220, 0.395] for Local Qwen3-4B. All four estimates divide by the same 628 A2 reference records. These are descriptive raw-field marginals; the paired inferential assembly analysis uses normalized recovery (Table S4).

### S5. Laboratory category support and schema scope

The laboratory annotation interface permits free-form category keys, whereas the output schema declares a closed 25-category set. All reference quintuples in the test set belong to this declared schema. Encoder, Cloud GPT-5.5, and Local Qwen3-4B generate 565, 0, and 20 off-schema quintuples, respectively. These outputs remain in the primary micro analysis. Category-projected diagnostic name counts are not additive because primary detection deduplicates test names across categories within each document.

Table S6 reports all 25 categories, including those absent from the reference data. Category-specific estimates are sensitive to small sample sizes: several categories contain only one to four quintuples, each concentrated in one case. Macro-F1 gives these categories the same weight as common categories, whereas micro-F1 pools counts across records. The secondary macro estimates are 0.112/0.404/0.425 for Encoder/Cloud/Local, with local minus cloud +0.021 [−0.025, +0.073].

**Table S6. Common-schema laboratory support and category-specific quintuple F1.**

| Declared category | Gold quintuples | Gold cases | Encoder | Cloud GPT-5.5 | Local Qwen3-4B |
| --- | --- | --- | --- | --- | --- |
| blood_test | 2006 | 187 | 0.663 | 0.941 | 0.964 |
| urinalysis | 1382 | 179 | 0.404 | 0.751 | 0.811 |
| biochem_blood_test | 1746 | 182 | 0.447 | 0.807 | 0.912 |
| coagulogram | 855 | 159 | 0.493 | 0.816 | 0.890 |
| syphilis_test | 112 | 108 | 0.000 | 0.860 | 0.873 |
| blood_electrolytes | 207 | 67 | 0.394 | 0.862 | 0.921 |
| fecal_analysis | 48 | 46 | 0.000 | 0.887 | 0.898 |
| cerebrospinal_fluid_analysis | 243 | 34 | 0.279 | 0.639 | 0.781 |
| blood_sugar_test | 89 | 39 | 0.119 | 0.434 | 0.695 |
| hiv_test | 18 | 14 | 0.000 | 0.571 | 0.634 |
| blood_type | 26 | 12 | 0.000 | 0.400 | 0.720 |
| hepatitis_test | 10 | 5 | 0.000 | 0.316 | 0.533 |
| hematocrit | 1 | 1 | 0.000 | 0.125 | 0.000 |
| sputum_analysis | 2 | 1 | 0.000 | 1.000 | 1.000 |
| malaria_test | 0 | 0 | 0.000 | 0.000 | 0.000 |
| arterial_blood_gas_test | 3 | 1 | 0.000 | 0.500 | 0.000 |
| alcohol_test | 1 | 1 | 0.000 | 0.000 | 0.000 |
| glycemic_profile | 4 | 1 | 0.000 | 0.190 | 0.000 |
| cholesterol_test | 0 | 0 | 0.000 | 0.000 | 0.000 |
| lipid_test | 0 | 0 | 0.000 | 0.000 | 0.000 |
| bacteriological_analysis | 0 | 0 | 0.000 | 0.000 | 0.000 |
| troponin_test | 0 | 0 | 0.000 | 0.000 | 0.000 |
| enterobius_eggs_test | 0 | 0 | 0.000 | 0.000 | 0.000 |
| gonococcal_test | 0 | 0 | 0.000 | 0.000 | 0.000 |
| rh_facto | 0 | 0 | 0.000 | 0.000 | 0.000 |

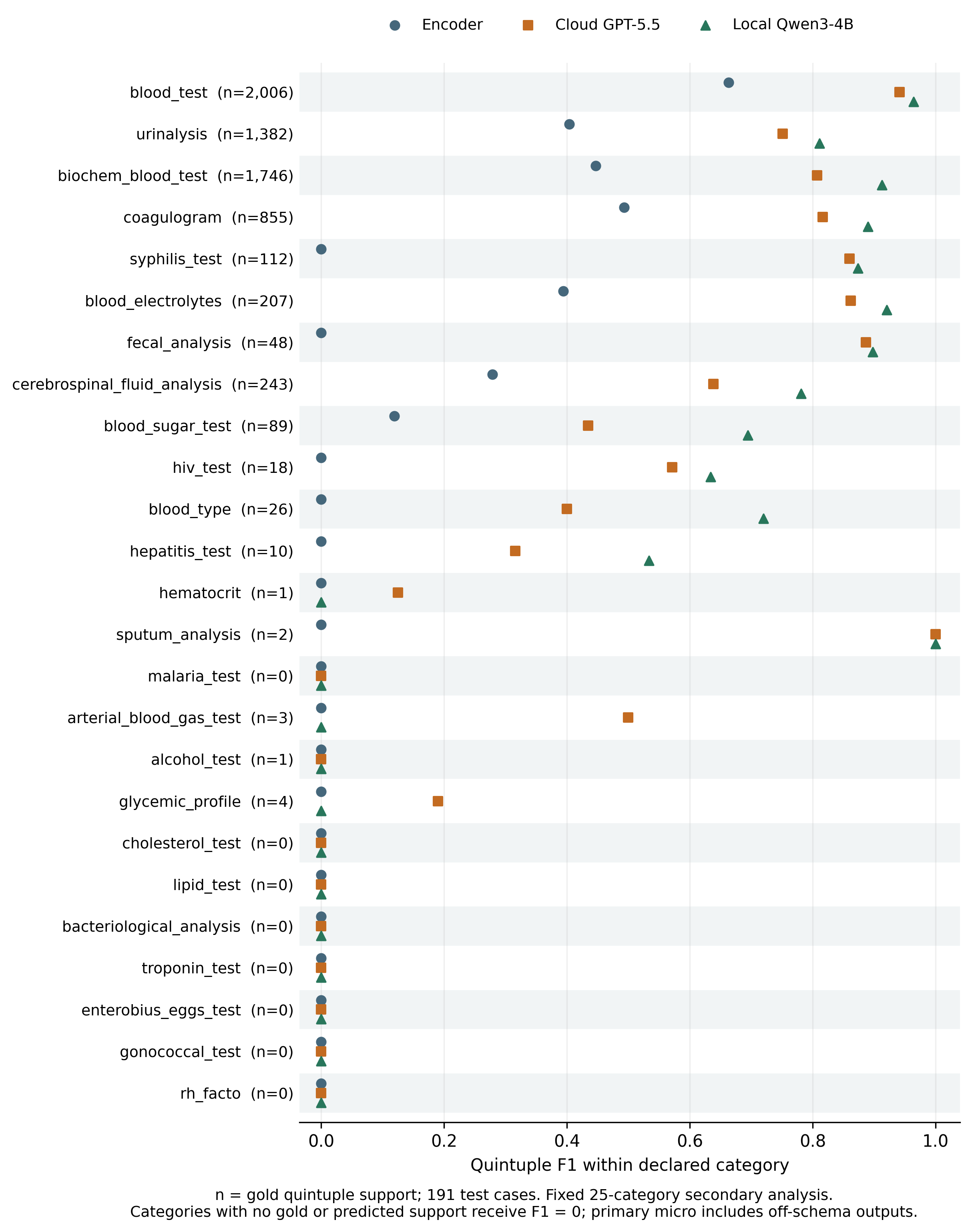

**Figure S1.** Laboratory quintuple F1 for all 25 declared categories on the 191-case test set. Parentheses give gold quintuple support; supporting-case counts are reported in Table S6. All arms use the same category set, with F1 zero for categories having neither gold nor predicted support. The literal schema label rh_facto is retained. This is a secondary schema-restricted analysis; off-schema outputs remain in primary micro scoring.

### S6. Curated-input controls, raw-document cascades, and corpus processing

The raw-document pipeline first extracts sections using the consensus-trained block encoder, with a laboratory keyword fallback, then applies the corresponding fine-tuned local record model. Medication single-window and all-block input construction are compared with curated-section controls using the same merged-model/vLLM stack. The main local medication benchmark uses a separate adapter inference path. Laboratory curated and cascade inputs are scored by the same test-name and quintuple projections.

The medication curated-control scores are drug-name F1 0.921 and normalized record recovery 0.377. The single-window cascade scores 0.752 and 0.204; the all-block cascade scores 0.779 and 0.246. Laboratory curated-control scores are 0.965 and 0.898, versus 0.917 and 0.811 in the cascade. All reference cases remain in the evaluation, including those with missing or unusable extraction outputs. Table S7 reports paired input-construction effects on the primary 149-case medication population and the original 191-case laboratory cohort. The laboratory cascade and its matched curated-input control retain the earlier model and cohort, which shares 149 reports with the revised laboratory benchmark.

**Table S7. Paired raw-document cascade-minus-curated-control effects.**

| Task | First minus control | Metric | Cases | Difference [95% CI] | Holm p |
| --- | --- | --- | --- | --- | --- |
| Medications | all blocks-curated control | Name F1 | 149 | -0.142 [-0.195, -0.095] | <0.001 |
| Medications | all blocks-curated control | Normalized record recovery | 149 | -0.131 [-0.180, -0.084] | <0.001 |
| Medications | single window-curated control | Name F1 | 149 | -0.169 [-0.214, -0.125] | <0.001 |
| Medications | single window-curated control | Normalized record recovery | 149 | -0.173 [-0.216, -0.131] | <0.001 |
| Laboratories | cascade-curated control | Name F1 | 191 | -0.048 [-0.065, -0.033] | <0.001 |
| Laboratories | cascade-curated control | Quintuple F1 | 191 | -0.087 [-0.111, -0.065] | <0.001 |

The corpus-scale run produced 2,540,326 medication records and 5,271,287 laboratory tuples in 13.3 H100 GPU-hours. Canonical reconciliation excludes 116 partition identifiers absent from the 193,101-case source population. The run includes 182,438 medication-section outputs and 191,983 laboratory-section outputs; laboratory sources comprise 178,449 encoder and 13,534 rule-based outputs. Of the outputs with sections found, 181,177 medication outputs and 187,615 laboratory outputs parse successfully. Section-found rates are 94.5% and 99.4%, and parseability among found-section outputs is 99.3% and 97.7%, respectively. Accuracy was assessed on the labelled benchmark and cascade cohorts described above.

An exploratory cascade-aware medication retraining experiment attained all-block detection F1 0.800 on the original medication cohort. This experiment used a separate model from the primary 149-case cascade comparison and the corpus-wide extraction run.

### S7. Task 1 development analyses

Task 1 development experiments compared output representations and model architectures using the original A1 annotations. Table S8 reports span macro-F1 from these experiments. The primary benchmark instead uses pooled micro-F1 against the adjudicated reference. Results are presented separately because the reference annotations, metrics, and adaptation settings differ.

**Table S8. Task 1 representation comparisons using original A1 annotations.**

| System | Adaptation | Original-gold span macro-F1 |
| --- | --- | --- |
| Qwen3-4B, plain block-ID | Zero-shot | 0.777 |
| Qwen3.6-27B, plain block-ID | Zero-shot | 0.481 |
| Qwen3.6-27B, richer block-ID prompt | Two demonstration examples; no fine-tuning | 0.778 |
| Qwen3-4B, block-ID | QLoRA, seed 42 | 0.781 |
| Qwen3-4B, block-ID | Full bf16 fine-tuning, seed 42 | 0.774 |
| Gazetteer | Rule-based | 0.212 |

Table S8 combines the original-annotation comparison report with the 332-case plain-prompt result. The two-example 27B condition uses few-shot prompting. Block-ID output represents sections as intervals over the document’s blocks. Token-length analyses found a median of 57 blocks per document and median output lengths of 85 block-ID tokens versus 762 free-form tokens.

Architecture ablations used a token-level encoder, seed 42, on the development set with original A1 annotations. Removing the CRF yielded F1 0.103; removing presence heads yielded 0.753; using the base encoder without domain-adaptive pretraining yielded 0.569. The corresponding full recipe scored approximately 0.72. These experiments preceded the final hierarchical block encoder. The examined self-training, gazetteer-feature, and ensemble variants provided no improvement in the tested configurations.

An input-length sensitivity analysis compared Task 1 local F1 0.9317 on all 332 cases with 0.9354 on the 329 cases whose text was not truncated. The restricted cohort excludes three long documents, so the difference reflects both input length and cohort composition.

### S8. Domain-adaptive pretraining exposure and temporal scope

Task 1 test text may have appeared in the encoder’s unsupervised masked-language-model pretraining without its task labels. Of 99,999 pretraining document identifiers, 99,936 map to the canonical corpus and 63 remain unmapped. All 332 Task 1 test cases map by exact identifiers: 179 are exposed and 153 are unexposed. The strata are not randomized and can differ in document composition.

**Table S9. Descriptive Task 1 pretraining-exposure strata.**

| Arm | Stratum | Cases | Pooled span F1 [95% CI] |
| --- | --- | --- | --- |
| Encoder | exposed | 179 | 0.919 [0.894, 0.942] |
| Encoder | unexposed | 153 | 0.919 [0.891, 0.944] |
| Cloud GPT-5.5 | exposed | 179 | 0.927 [0.905, 0.949] |
| Cloud GPT-5.5 | unexposed | 153 | 0.926 [0.898, 0.952] |
| Local Qwen3-4B | exposed | 179 | 0.930 [0.907, 0.950] |
| Local Qwen3-4B | unexposed | 153 | 0.934 [0.907, 0.956] |

Encoder F1 rounds to 0.919 in both exposure groups. Because group membership is non-random, this comparison cannot separate pretraining exposure from differences in document composition. Scores for the cloud and local generative models are reported on the same groups to describe performance across those documents. The evaluation uses internal splits of 2013–2019 records with 2026-era generative models; performance on later documentation, external institutions, and other specialties remains untested. Language-specific performance was not established.

### S9. Qualitative error classes

Table S10 summarizes error classes documented in the qualitative analysis. Synthetic examples illustrate each class without reproducing patient text. The analysis describes types of disagreement and extraction error; class prevalence was not quantified.

**Table S10. Qualitative error classes with synthetic illustrations.**

| Class | Synthetic illustration | Interpretation and observed family |
| --- | --- | --- |
| Dosage–strength slot assignment | Reference strength=5 мг, output dosage=5 мг | Same surface quantity in a different slot fails assembly; observed across families. |
| Form/route vocabulary | Reference form=solution, output form=р-р | Resolved codes may agree despite raw strings differing; documented for Cloud. |
| Spurious fragment detection | A stray dose token such as 200 is output as a drug name | A detection false positive; documented for Encoder. |
| Dense medication list omissions | Several infusion drugs occur in one densely punctuated line; one is omitted | An unmatched reference record fails detection and recovery; documented for Local. |
| Medication scope | A remote home-medication mention is included in a task-specific hospital list | Scope error despite a plausible drug name; documented for Cloud. |
| Drug-name trimming | Reference Brand A (substance B), output Brand A | May pass name matching but fail raw-name equality required for assembly; documented for Local. |
| Frequency surface form | Reference 1тх2р, output 2 раза в день | Both may encode two daily doses but differ in tablets per dose, which remains relevant after normalization; documented for Local. |
| Section boundary granularity | Two adjacent laboratory subsections are merged into one interval | Same-label content can differ in span partitioning and IoU; observed across families. |

### S10. Prompts, model settings, and reproducibility

#### Experimental model settings

The cloud evaluations used openai/gpt-5.5 through OpenRouter in August 2026. The local generative family is Qwen3-4B, adapted separately for each task. The encoder uses multilingual ModernBERT with domain-adaptive masked-language-model pretraining and task-specific structured prediction heads. Table S11 summarizes the documented experimental settings.

**Table S11. Documented experimental model settings.**

| Component | Documented settings |
| --- | --- |
| Encoder, Task 1 | Block embeddings; two-layer bidirectional LSTM, 256 hidden units per direction; BIO head and linear-chain CRF; eight epochs; batch 1 with accumulation 8; validation-based selection |
| Local Qwen3-4B, Task 1 | Four-bit NF4 QLoRA, rank 16, alpha 32, all-linear targets; three epochs; completion-only loss; 8,192-token context; greedy generation |
| Cloud GPT-5.5, Task 1 | OpenRouter chat-completion API; section runner specifies temperature 0 and max_tokens 2,048; section instruction reproduced below |
| Local Qwen3-4B, medication task | Task-specific bf16 LoRA/RSLoRA adaptation; structured record outputs; greedy inference |
| Local Qwen3-4B, laboratory task | Qwen/Qwen3-4B-Instruct-2507; four-bit QLoRA; rank 32, alpha 64, dropout 0.05; RSLoRA targeting all seven attention/MLP projections; five epochs; learning rate 1.5e-4 with cosine schedule; effective batch 16; best checkpoint by validation quintuple F1; greedy decoding, temperature 0, max_new_tokens 4,096; no constrained decoding |
| Medication merged-model cascade/control | vLLM, bfloat16; model context 8,192; temperature 0; output cap 3,400; source implementation applies a context-budget guard; controls and cascades share this stack, while the main benchmark uses a separate adapter path |
| Cloud GPT-5.5, laboratory task | openai/gpt-5.5 through OpenRouter; zero-shot; temperature 0; response_format=json_object; default reasoning effort; dictionary-based system prompt with an annotation-guide addendum |
| Cloud GPT-5.5, medication task | Zero-shot structured medication extraction |

The following prompts reproduce the task instructions, with formatting tokens preserved. Clinical inputs are represented by template variables or synthetic examples. The complete cloud laboratory system prompt and user template are supplied separately in cloud_lab_prompt.txt.

#### Task 1 cloud section prompt

System message:

You are a helpful assistant for medical document segmentation.

The user message consists of the following instruction immediately followed by the numbered document blocks:

Ты — ассистент по сегментации медицинских документов для русскоязычных выписок. Текст выписки дан в виде пронумерованных блоков [B0001], [B0002], … Задача: указать интервалы блоков для ТРЁХ разделов:

1. lab_results (результаты лабораторных исследований) Маркеры: «Лабораторные анализы», «ОАК», «биохимия», «анализ крови», гемоглобин, лейкоциты, СОЭ, тромбоциты, глюкоза, холестерин, АЛТ, АСТ, билирубин, креатинин, мочевина
2. prescription (препараты, рекомендованные ПРИ ВЫПИСКЕ пациенту домой) Маркеры: «при выписке», «дома», «продолжить», «рекомендовано», «лечение после выписки», список препаратов с дозировкой и длительностью приёма ВАЖНО: это препараты для ДОМАШНЕГО приёма, а не стационарного!
3. drugs_hospital (препараты, назначенные В СТАЦИОНАРЕ во время госпитализации) Маркеры: «Лечение», «в/в», «в/м», «капельно», «№ инъекций», «схема лечения», «проведённая терапия», назначения с путём введения (внутривенно, внутримышечно) ВАЖНО: это препараты, которые пациент получал В БОЛЬНИЦЕ!

Формат ответа: JSON-массив объектов {“first”: “B0007”, “last”: “B0012”, “label”: “”}. Один раздел может состоять из нескольких интервалов. Если раздела нет — не включай его. Верни ТОЛЬКО JSON.

A non-clinical synthetic illustration of the appended representation is:

[B0001] Лабораторные анализы
[B0002] Глюкоза 5,0 ммоль/л
[B0003] Рекомендации при выписке
[B0004] Препарат А 5 мг утром

An illustrative output for the laboratory section is [{"first":"B0001","last":"B0002","label":"lab_results"}]. This synthetic example shows the Task 1 output format.

#### Local medication prompt

The retained local medication fine-tuning formatter and merged-model cascade implementation contain the same system instruction:

Вы — модель структурированного извлечения информации о лекарственных препаратах из медицинских выписок.

ЗАДАЧА: разобрать текст с лекарствами на отдельные записи. Для каждого препарата извлечь поля: - drug_name: название препарата (ОБЯЗАТЕЛЬНО — всегда заполнять) - dosage: количество/объём (например: 5,0 / 200,0 / 10мл) - strength: концентрация или единичная доза (например: 25% / 10мг / 400мг) - form: лекарственная форма (например: табл / капс / р-р / амп) - route: путь введения (например: в/в / в/м / п/к / перорально) - frequency: кратность приёма (например: 1тх3р / 2 раза в день / утром / на ночь) - duration: длительность или курс (например: №10 / 1 мес / длительно / постоянно)

ПРАВИЛА: 1. Если поле НЕ указано в тексте — ставьте null (не придумывайте значения). 2. Копируйте значения ДОСЛОВНО из текста, не нормализуйте и не переводите. 3. Не объединяйте несколько препаратов в одну запись. 4. Отвечайте ТОЛЬКО валидным JSON, без Markdown и пояснений.

User template ({text} is the supplied medication section; doubled braces in the source template render literal JSON braces):

Разберите следующий текст с лекарствами на структурированные записи.

Текст: {text}

Верните JSON: {“medications”: […]}

The local formatter surrounds the system and user strings with <|im_start|>system, <|im_end|>, <|im_start|>user, and an open <|im_start|>assistant generation prefix. The explicit instruction not to normalize describes generation; deterministic normalization is a subsequent scoring operation.

#### Cloud laboratory prompt

The cloud laboratory system prompt combines dictionaries for category and parameter names, units, and qualitative values with an annotation-guide addendum. It requests a JSON object nested by category and date, containing test, value, and unit fields. The addendum specifies date inheritance, category inference from context, typical-unit inference, and corrections for implausible value–unit combinations. It also extends the dictionaries with additional parameters and categories. These instructions guide generation; the scoring procedures are described in S2.

The complete system message, including all dictionaries, reference tables, and guide rules, is provided in the accompanying supplementary prompt file, <cloud_lab_prompt.txt>. Its user message uses the following template:

Return output as JSON. Extract laboratory test results from the following medical text:

{text}

#### Local laboratory cascade prompt

System message:

You are a medical data extraction assistant. Your task is to extract structured laboratory test results from unstructured medical texts. Extract all test names, values, units, and dates, organizing them by test category (blood tests, urinalysis, biochemistry, etc.).

User template:

Extract laboratory test results from the following medical text:

{text}

The prompt above was used by the local laboratory cascade. Table S6 specifies the 25-category scoring schema.

#### Access and computational reproducibility

Evaluation records retain document-level sufficient counts and source hashes linking code, normalization vocabularies, predictions, annotations, and cohort definitions. The plotting script uses aggregate estimates and category support to render Figures 1, 3, and S1. Its CSV outputs contain aggregate differences, confidence intervals, category support counts, and source-file hashes. Clinical text and case identifiers are excluded from these public plotting assets.

The evaluation-code supplement contains scoring and statistical source, normalization dictionaries, and a synthetic-count example. Clinical documents, annotations, predictions, and trained weights are excluded. Empirical reproduction requires these study data and model artifacts in addition to the code. Access to the anonymized clinical corpus may be granted under an institutional agreement, subject to approval.

### S11. Reporting guideline mapping

Table S12 maps the 19 TRIPOD-LLM items to the relevant sections of this extraction study and identifies partially reported or inapplicable elements.[1] The study evaluates structured extraction retrospectively, so the map distinguishes extraction performance from patient-risk calibration and prospective clinical implementation.

**Table S12. TRIPOD-LLM reporting map and journal-specific applicability notes.**

| Item(s) | Topic | Status | Manuscript/supplement location and limitation |
| --- | --- | --- | --- |
| 1–2 | Title and abstract | Addressed | Main title and structured Abstract state the extraction tasks, model comparison, retrospective setting, and primary findings. |
| 3–4 | Background and objectives | Addressed | Main Introduction motivates health-system extraction and task-dependent model selection. |
| 5a–e | Data | Partial | Main Study design, data, and cohorts; S1 and S8 describe source, dates, task populations, identity exclusions and pretraining exposure. External/temporal validation and complete patient-level linkage are not established. |
| 6a–e | Methods | Partial | Main Task-specific representations and model families; Outcomes and statistical analysis; S2 and S10 describe the model architectures, experimental settings, and evaluation procedures. |
| 7a–e | Output | Addressed for extraction | Main Outcomes and S2 define section spans, seven-field records, laboratory names/quintuples, normalization, unmatched records, and schema scope. |
| 8a–c | Annotation | Addressed for reported protocol | Main Annotation and human reference; S1 and S3 identify protocol roles, common-A2 pairing, and adjudication by a third MD resident. Human agreement uses the original annotations before adjudication. |
| 9a–b | Prompting | Partial | S10 reproduces cloud section and local medication/laboratory prompts and describes the cloud laboratory prompt. The complete cloud laboratory system message, dictionaries, annotation-guide addendum, and user template are supplied in cloud_lab_prompt.txt. |
| 10 | Summarization | Not applicable | The outputs are section intervals and structured records. |
| 11 | Fine-tuning | Partial | Main model-family descriptions and S10 report task-specific adaptation settings; S7 describes development ablations. |
| 12 | Computational resources | Partial | Main raw-document system evaluation and corpus-processing results; S6 and S10. Resource reporting covers GPU runtime and selected API charges; energy and lifecycle costs were not measured. |
| 13 | Ethics | Partial | Main Statements and Declarations report ethics approval and consent waiver. Main Methods describe cloud-input preprocessing; data access is restricted. |
| 14a–f | Open science | Partial | Main Material and code availability and S10 describe the evaluation-code supplement, synthetic example, aggregate plotting assets, and institutional data-access arrangements. |
| 15 | Patient and public involvement | N/A | Patient and public were not involved. |
| 16a–d | Participants/data flow | Partial | Main cohorts and Figure 1; S1 reports cohort counts and exclusions. Full demographic or site-stratified representativeness analyses are unavailable. |
| 17 | Performance | Addressed for defined endpoints | Main Tables 1–3 and S2–S6 report pooled metrics, denominators, marginal intervals, paired comparisons, sensitivities, and human comparisons. |
| 18 | Model updating | Partial | S6 describes an exploratory cascade-aware retraining experiment using a separate model from the primary benchmark and corpus run. Prospective updating was outside of scope. |
| 19a–g | Discussion | Partial | Main Discussion and Conclusion; S7–S10 qualify historical analyses, conventions, cohort, exposure, transfer, and reproducibility. Clinical benefits and deployment effectiveness remain untested. |
| Journal-specific | Calibration and clinical implementation | Risk calibration not applicable; clinical implementation not evaluated | Outputs are structured records. Live EHR integration and prospective patient outcomes were outside the evaluation scope. |
| Journal-specific | AI-use disclosure | Partial | Main Methods describe the extraction models; Use of artificial intelligence discloses copyediting and analytic assistance and identifies the documented drafting tool. |
